# Canine olfaction combined with Bayesian modeling for multi-cancer detection from breath samples: a Phase-2 study in India

**DOI:** 10.1101/2025.09.21.25336259

**Authors:** Sanjeev Kulgod, Basavaraj R. Patil, Shashidhar K, Rakesh S Ramesh, Kiran Kulkarni, Somashekhar S P, Swaratika Majumdar, Akshita Singh, Claire Guest, Rob Harris, Sahana Shanbhag, Ido Aviram, Itamar Bitan, Akash Kulgod

## Abstract

**PURPOSE:** Low-cost, acceptable, and high-sensitivity triage tests are needed to address the challenge of low cancer prevalence in population screening, particularly in low- and middle-income countries (LMICs). Breath-based canine olfaction has the potential to serve this role; however, evidence to date has been mostly limited to high-income countries and relatively small, single-cancer studies. We evaluated the analytical validity of a multi-cancer breath detection system using trained dogs and Bayesian fusion modeling.

**PATIENTS AND METHODS:** We conducted an assessor-masked, multi-center case–control study across six hospitals in Karnataka, India (March 2024–June 2025; CTRI/2024/10/075938). A total of 3,275 participants were enrolled: 1,773 for training and 1,502 for testing. The test cohort comprised 283 treatment-naive, biopsy-confirmed cancer patients (seven major cancer groups) and 1,219 controls (healthy, non-oncologic chronic disease, and benign biopsy). Breath was collected on cotton masks, stored under −20°C cold-chain conditions, and presented on a sniffing platform to trained detection dogs. Individual responses were integrated with Bayesian fusion incorporating historical dog performance and participant-level sample variables.

**RESULTS:** The fusion system achieved 91.5% sensitivity (95% CI, 88.0 – 94.8) and 90.8% specificity (95% CI, 89.1 – 92.5), with an area under the ROC curve (AUC) of 0.962 (95% CI, 0.951 - 0.971). Sensitivity was 89.6% in early-stages (Stage I–II), and was relatively consistent across major cancer types.

**CONCLUSION:** In a 1,502-participant test cohort, canine olfaction–Bayesian fusion achieved high accuracy for multi-cancer detection from breath, with stable performance across stages. These data establish the analytical validity and support the prospective evaluation of true screening populations.

**Context Summary:** *Key Objective:* Does canine olfaction combined with Bayesian modeling maintain analytical validity for multi-cancer breath screening in a large assessor-masked study in India?

*Knowledge Generated:* Detection dogs achieved sensitivity and specificity above 90% (AUC 0.962) in 1502 participants, with comparable performance across early- and late-stage cancers.

## Introduction

Multi-cancer early detection (MCED) tests^1, 2^ aim to address the persistent problem of late-stage cancer diagnosis, which lowers treatment success and contributes substantially to global cancer mortality (9.7 million deaths in 2022).^3^ Despite their promise, current blood-based MCED approaches have limited sensitivity in early-stage disease^4^ and require costly sequencing infrastructure, trained personnel, and cold-chain logistics, with per-test costs of approximately US$500-1,000.^5^ These characteristics constrain population-scale deployment, especially in low- and middle-income countries (LMICs).

India, an LMIC with a population exceeding 1.4 billion, faces rising cancer incidence and, for some malignancies (e.g., breast cancer), an earlier age at onset than high-income settings.^6^ Screening coverage remains low (approximately 1% ever screened), and a high proportion of cancers are detected at stages III–IV (approximately 80%), underscoring the need for accessible and acceptable screening alternatives.^7^ At the same time, India’s sheer population size and relatively lower age-standardized incidence compared with many high-income countries^3^ make the problem of low prevalence in asymptomatic cohorts particularly acute. Even tests with very high specificity will generate large absolute numbers of false positives relative to true cancers, thus straining the already limited diagnostic resources.

A central challenge for early detection is therefore statistical as well as logistical: in low-prevalence populations, even highly specific tests yield modest positive predictive value (PPV).^8^ The CanTest Framework was developed to guide evaluation of new tests across the diagnostic pathway, emphasizing the need for inexpensive, acceptable, high-sensitivity “triage” tests that enrich pretest probability so that confirmatory diagnostics can be deployed more efficiently.^9^ This logic is especially pertinent in LMICs such as India, where confirmatory investigations are costly, invasive, and concentrated in urban centers, and patient acceptance of invasive procedures is low. Against this backdrop, breath-based canine detection could serve as a first-layer triage: non-invasive, low-cost, and operationally simple, it could filter large populations so that confirmatory diagnostics are focused on a smaller, higher-risk subset.^10^

Over two decades of studies have indicated that trained canines can detect cancer-associated volatile organic compounds (VOCs) emitted from blood, urine, saliva, sweat, feces, and breath.^11–13^ Breath is especially attractive, literature reviews have catalogued over 1,400 distinct VOCs in human breath, capturing tumor-related metabolic signatures.^14^ Prior studies have evaluated canine detection for single cancers (e.g., breast^15^ and lung^16, 17^) and, more recently, multi-cancer settings,^18^ often reporting high sensitivity and specificity. Because breath collection is non-invasive and has modest storage/transport needs relative to phlebotomy, and canine teams can be trained and maintained at a comparatively low cost, breath sampling may support scalable population-level multicancer triage, enabling more effective use of organ-specific screening resources.

This study covers a Phase-2^9^ evaluation of trained dogs for multi-cancer detection from breath collected on face masks in India. We conducted an assessor-masked, multi-center case–control evaluation to estimate accuracy across cancer types and stages and assessed whether a Bayesian fusion integrating individual-dog historical performance, participant/sample covariates, and sensor-derived behavioral signals improves accuracy, stability, and reproducibility relative to individual dog reads.

## Methods

### Participants

We conducted an assessor-masked, multi-center case–control diagnostic accuracy study at six hospitals in Karnataka, India : three in Hubballi (RadOn Cancer Center, Karnataka Cancer Therapy and Research Institute, Karnataka Medical College & Research Institute) and three in Bengaluru (St John’s Medical College, Narayana Health City, Aster CMI). The two Hubballi public/tertiary centers draw predominantly rural catchment populations, whereas one Hubballi private center and the Bengaluru centers serve largely urban/semi-urban populations, yielding broad demographic and exposure diversity. The study was registered under the Clinical Trials Registry of India (CTRI/2024/10/075938)

Recruitment occurred across oncology and non-oncology services (e.g,, internal medicine, surgery, gastroenterology and gynecology). Case enrollment was opportunistic with no restriction on the primary tumor site (multi-cancer intent); principal investigators at each site, supported by clinical research coordinators, approached eligible patients in outpatient clinics, inpatient wards, and procedure areas. Recruitment for control participants also occurred at community screening camps conducted by partner hospitals.

*The patients* were adults (*>*18 years old) with biopsy/FNAC-confirmed malignancy and no prior cancer. Breath collection was performed before treatment to avoid confounding from acute procedures or therapy. The majority of cases were recruited after confirmation of the diagnosis, although a small minority were collected before biopsy.

*The controls* were also adults (*>*18 years) and comprised (i) healthy volunteers, (ii) patients with non-oncologic chronic conditions, and (iii) patients with benign biopsy results. To reduce setting-related confounding, where feasible, controls from hospitals were recruited in the same rooms as cases within the same clinical departments. Key exclusions for all participants were pregnancy/postpartum *<* 90 days and any communicable respiratory illness, the latter to ensure the bio-safety of all team members.

All participants provided written informed consent in their preferred language (Kannada/Hindi/English). Site-specific ethics approvals was obtained from the institutional committees; and the study procedures adhered to the Declaration of Helsinki and ICH-GCP. For transparency, the protocol was registered with the Clinical Trials Registry of India in October 2024 (CTRI/2024/03/061847). Demographics, tobacco use (smoking/chewing), and comorbidities were recorded for all participants. For cases, tumor site, histology, and stage were extracted from the medical records. The dataset was prospectively partitioned into disjoint training and testing sets at the participant level and all samples used for testing were novel samples from novel individuals at the time of presentation to the dogs.

### Index Test and Procedures

Exhaled breath volatile organic compounds (VOCs) collected on cotton face masks were evaluated by trained detection dogs using a standardized indication protocol. The primary endpoint was cancer vs non-cancer classification for each sample. For all tests, samples were identified only by QR codes; clinical data and reference results were withheld from the handler logging the canine indications. The *reference standard* for case/control status was histopathology or cytology from routine care (biopsy/FNAC); imaging was used for the clinical context only, and index-test results did not influence clinical management.

### Sample Collection (Pre-analytical Conditions)

The participants were instructed to perform 10 minutes of tidal breathing wearing a standardized cotton surgical mask. Trained research coordinators handled masks with gloves to minimize contamination, immediately sealed them in aluminum barrier bags, and logged unique QR codes under chain-of-custody. All participants in this study provided only one sample. Short-term storage was at 4-8^°^C (typically *<*24 hours during on-site collection), followed by site-storage for 3 weeks at −20^°^C, and eventually transferred to the central testing laboratory at −20^°^C. Collection procedures were identical for cases and controls and, where feasible, performed in the same clinical rooms to limit location- or workflow-related confounding. The samples were stored for up to six months before testing.

### Canine Team and Experimental Platform

Seven adult dogs (four Beagles, one Labrador, one Labrador-Indie mix, and one Dutch Shepherd-Belgian Malinois mix; 6 female, 1 male; ages 1-3 years) underwent positive-reinforcement training (10 weeks preceding each of the two trial sessions) at Dognosis facilities. Dogs were housed with daily exercise, veterinary oversight, and environmental enrichment, meeting the Five Freedoms and adhering to international welfare frameworks.^19, 20^ The protocol was approved by the Vigilomics Institutional Animal Ethics Committee (IAEC) under CCSEA registration and complied with the Prevention of Cruelty to Animals Act (1960) and CCSEA guidelines.

Training used 1,773 samples from unique participants (373 cancer, 1,400 control); a sample was not used more than three times during training, and the same samples were not used in the training and testing sets. Both training and testing employed a custom eight-port sniffing workstation equipped with synchronized video and infrared sensors for automated timing and behavioral capture, as described in detail elsewhere.^21^ QR scanners automated sample identification and logging, barrier gates controlled access timing, an automated feeder delivered rewards. The sensor streams were synchronized using software.^22^

### Testing Procedure

The testing sessions were pre-declared and logged daily. Arrays of eight samples were randomly selected from the testing pool, yielding 40-70 samples per day. The dogs were evaluated individually in sequential runs. Two trainers and two laboratory technicians conducted the sessions; the handler calling indications remained masked to the sample identity throughout. Each sample was independently assessed by a minimum of three dogs using a pre-specified indication protocol (binary alert vs pass). Trials affected by technical interruptions (e.g., hardware interruption) or discontinued by the handler due to a lack of canine motivation to work were re-run when feasible or excluded from the primary accuracy analysis.

### Bayesian Fusion Algorithm and Statistical Analysis

Individual dog indications and sample/demographic co-variates were combined into a Bayesian fusion framework. Dog-specific likelihoods were estimated from training folds (historical sensitivity/specificity per dog), and *co-variate informed prior odds* were obtained from a logistic model fit on the training set. For a given sample 𝔖, let *C* ∈ {0, 1} denote the cancer status; Prior *P*_0_ = *P* (*C* = 1 | *x*_𝔖_), [See Priors]. Dogs *i* = 1, …, *D* produce indications *I*_*i*_ ∈ {0, 1} when present; each dog has sensitivity *s*_*i*_ and specificity *c*_*i*_ estimated from the training confusion matrices. For the presented dogs *i* ∈ 𝔻 (𝔖), using per-dog sensitivity *s*_*i*_ and specificity *c*_*i*_:

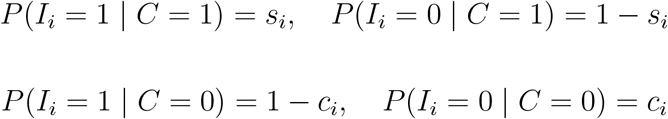

Assuming conditional independence across dogs, the likelihoods are:

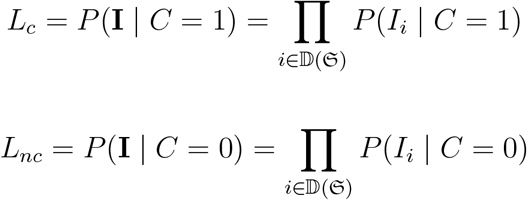

Optionally tempering the likelihoods with *τ* ≥ 0:

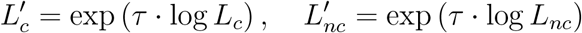

The Bayesian update with the prior *P*_0_ is:

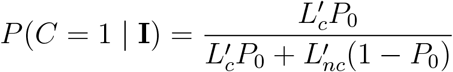

Posterior probabilities were compared with varying decision thresholds; threshold–performance trade-offs are shown as ROC/operating-point analyses. All fusion computations were performed on a test set without re-estimation of the dog performance parameters.

The primary accuracy measures were sensitivity, specificity, and area under the ROC curve (AUC) for the testing set with two-sided 95% CIs. Pre-specified stratification included stage (I–II vs III–IV), cancer type, sex, age, tobacco use, tumor site, and dog identity.

### Prior Prediction Model

#### Feature Engineering and Preprocessing

Engineered variables included age, sex, tobacco exposure, family history of cancer, self-reported persistent symptoms, and other variables. Numeric variables were imputed and standardized, while categorical variables are imputed and one-hot encoded. For sample *x*_𝔖_, let the features be *ϕ*(*x*_𝔖_)

#### Classifier

A supervised classifier (logistic regression) was trained on training historical data to produce *P*_0_(*x*) = *P* (*C*=1 | *x*_𝔖_). For the logistic regression,

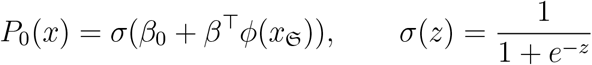

where *ϕ*(*x*_𝔖_) is the feature vector of engineered covariates for sample *x*_𝔖_, *β* is the vector of regression coefficients, and *β*_0_ is the intercept term.

## Results

### Participants

A total of 3,625 individuals were assessed across the six hospitals for eligibility. After 350 exclusions (age, communicable respiratory disease, and pregnancy); 3,275 participants were enrolled. Of these, 1,773 contributed to the training cohort and 1,502 to the testing cohort. Within the control group in the test set, participants were comprised of healthy volunteers (n = 789), patients with chronic non-malignant conditions (n = 426), and individuals with benign biopsy findings (n = 4).

The final test cohort comprised 283 biopsy-confirmed cancers and 1,219 controls, with a STARD flow diagram summarizing the recruitment (Figure 1).

**Figure 1.**
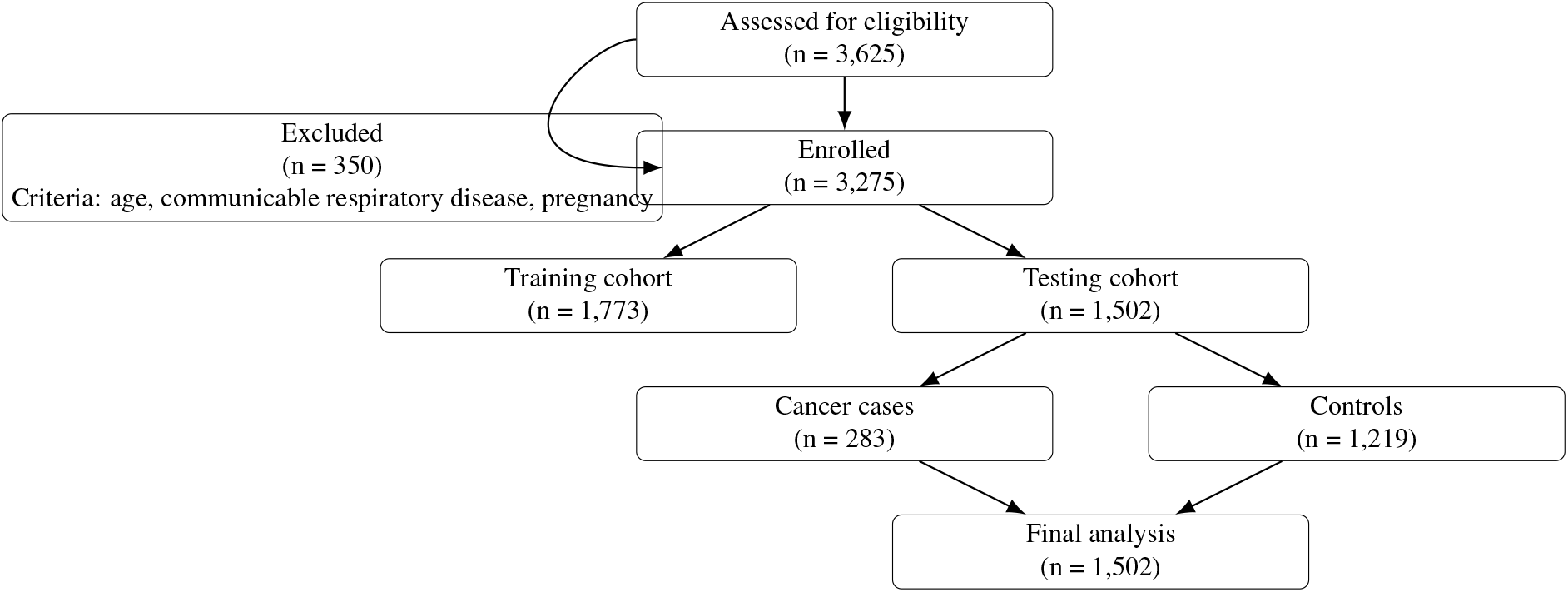
STARD flow diagram of participant disposition showing exclusions, training allocation, and test cohort composition.

Cases tended to be older than controls (mean 55.3 vs 48.5 years) and had higher prevalence tobacco use, particularly chewing (38.2% vs 14.9%). Smoking history was also more common among the cases (14.1% vs 7.6%). However, given the larger number of controls overall (n = 1,219), the absolute number of tobacco-exposed individuals was comparable between the groups (e.g., 182 controls with chewing history vs 108 cases). This balance reduces the likelihood of canine discrimination reflecting tobacco exposure. Hypertension and diabetes were similarly distributed among the groups (Table 1). No adverse events occurred during breath collection or testing.

**Table 1:**
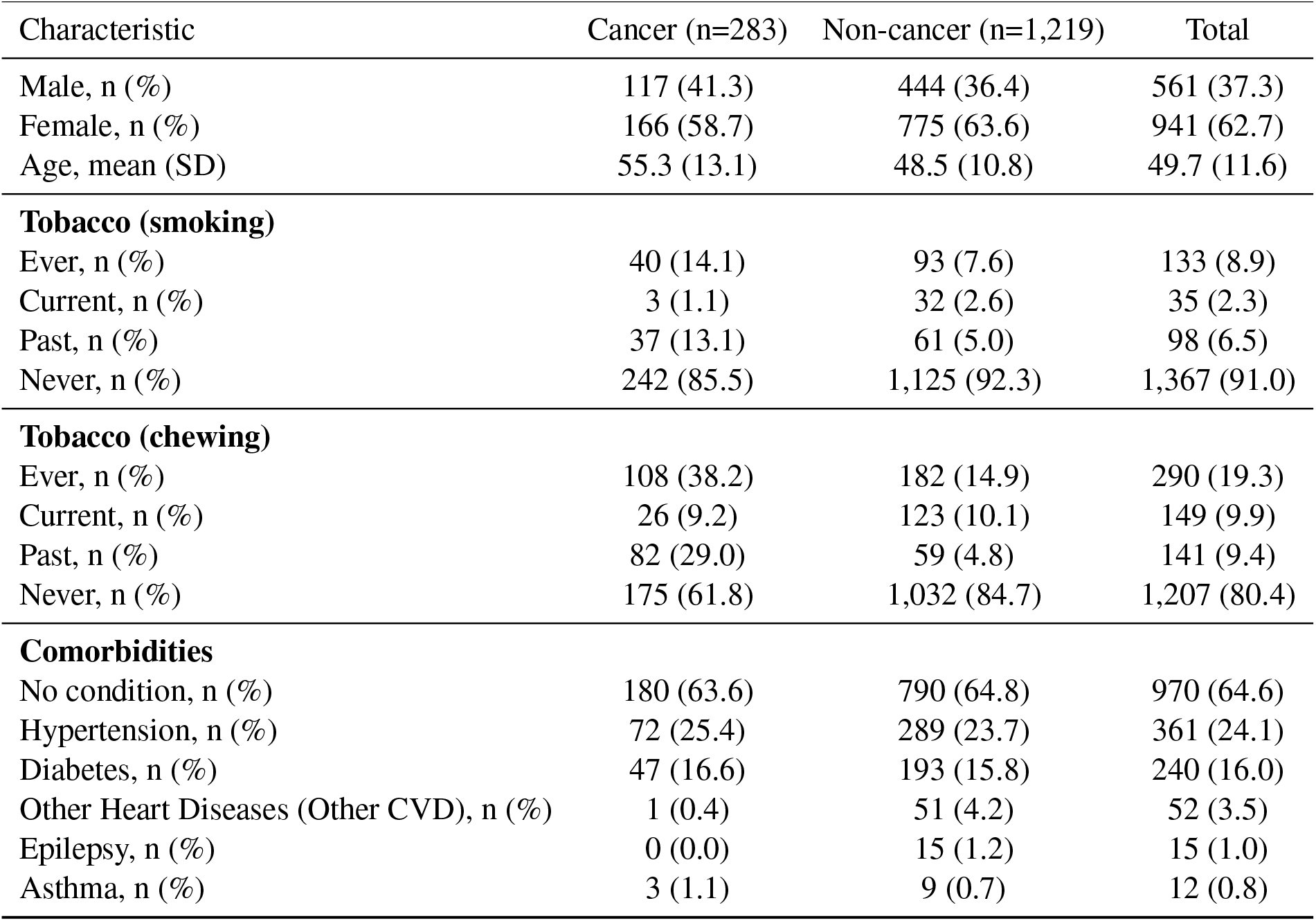
Baseline demographic and clinical characteristics of the test cohort (n = 1,502)

### Cancer distribution

The distribution of the cancer groups in the test cohort (n = 284 cancer diagnoses from 283 participants) is summarized in Table 2. One participant had two distinct primary cancers, both of which were included in the overall count but only one of them (Breast cancer) was used to determine performance. Grouping rules (site and histology mapping) are detailed in Supplementary Methods S2, with the full ICD-10/ICD-O mapping in Table S1. Head and neck cancers accounted for nearly one-third of the cases, consistent with the high prevalence of tobacco-related oral malignancies in India. Upper gastrointestinal and breast cancers together contributed to another 40% of cases, reflecting the dual burden of lifestyle-associated and hormonally driven malignancies in the region. Gynecological cancers were also well represented, while lung and male genitourinary cancers were fewer in number, paralleling the local incidence trends.

**Table 2:** Cancer-group distribution in the test cohort (n=284)

| Cancer group | N | % of cancers |
| --- | --- | --- |
| Head and Neck | 88 | 31 |
| Breast Cancer | 48 | 16.9 |
| Lung and Thoracic | 8 | 2.8 |
| Upper GI | 59 | 20.8 |
| Lower GI | 23 | 8.1 |
| Gynecological | 40 | 14.1 |
| Genitourinary (male) | 4 | 1.4 |
| Other | 14 | 4.9 |
| Total | 284 | 100.0 |

### Primary accuracy

Bayesian fusion achieved an overall 91.5% sensitivity (95% CI, 88.0 – 94.8) and 90.8% specificity (95% CI, 89.1 – 92.5), with an area under the ROC curve (AUC) of 0.962 (95% CI, 0.951 - 0.971) (Figure 2). At a probability threshold of 0.5, which was used as the default operating point for binary classification, the model correctly identified 259 of 283 cancers and 1107 of 1,219 controls, with 24 false negatives and 112 false positives (Table 3). As expected, alternative thresholds yielded predictable trade-offs between sensitivity and specificity, captured in the ROC profile; full sensitivity/specificity curves across thresholds are shown in Figure 3). Positive and negative predictive values are not reported given the case–control design and artificially inflated prevalence, but will require evaluation in prospective screening populations.

**Table 3:** Confusion matrix, test set.

| Table 3: Confusion matrix, test set |  |  |
| --- | --- | --- |
| Fusion prediction | Reference (biopsy/FNAC) |  |
|  | Cancer (n=283) | Non-cancer (n=1,219) |
| Positive | **TP = 259** | **FP = 112** |
| Negative | **FN = 24** | **TN = 1107** |

**Figure 2.**
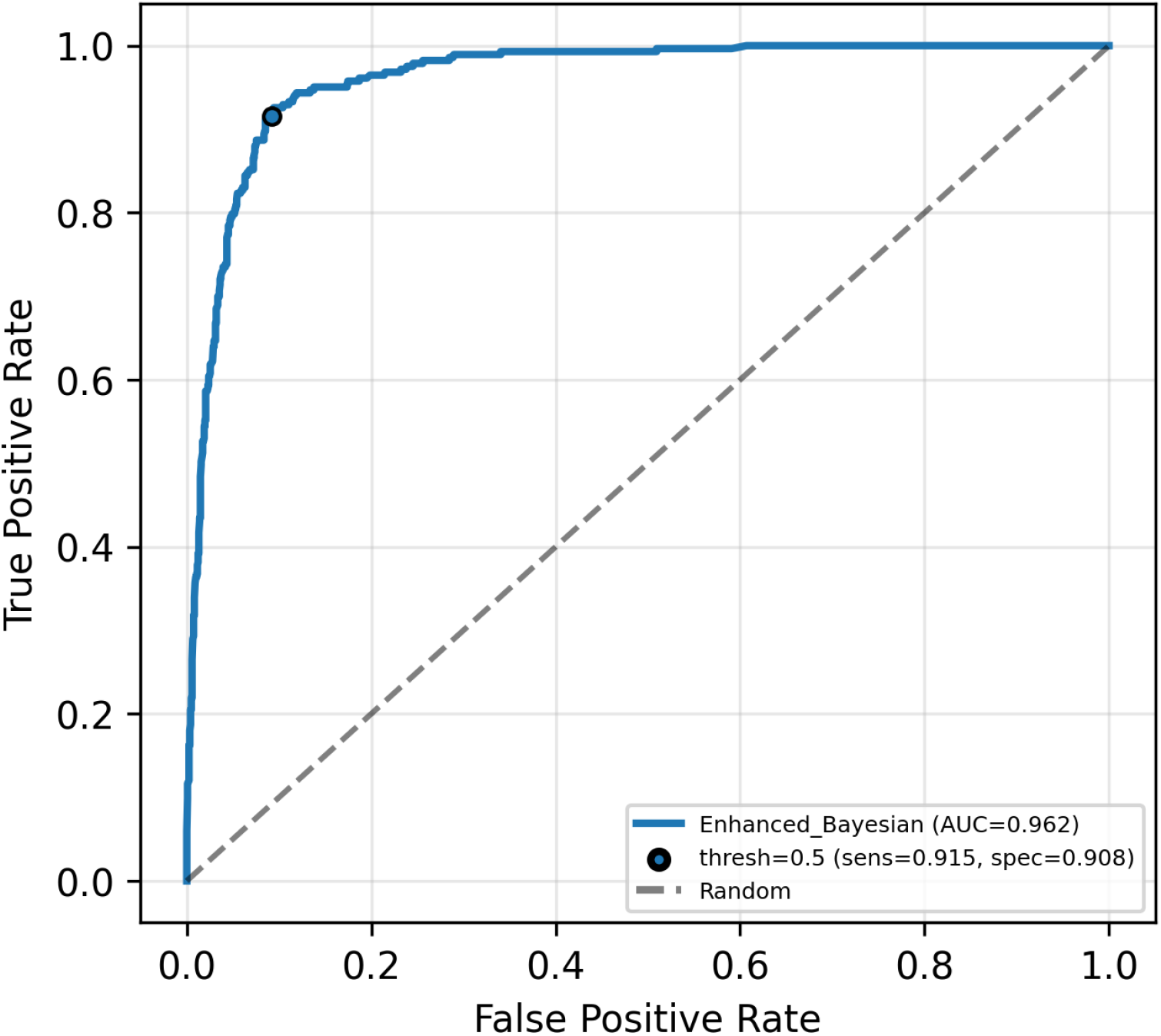
ROC curve for the Bayesian fusion (AUC = 0.962)

**Figure 3.**
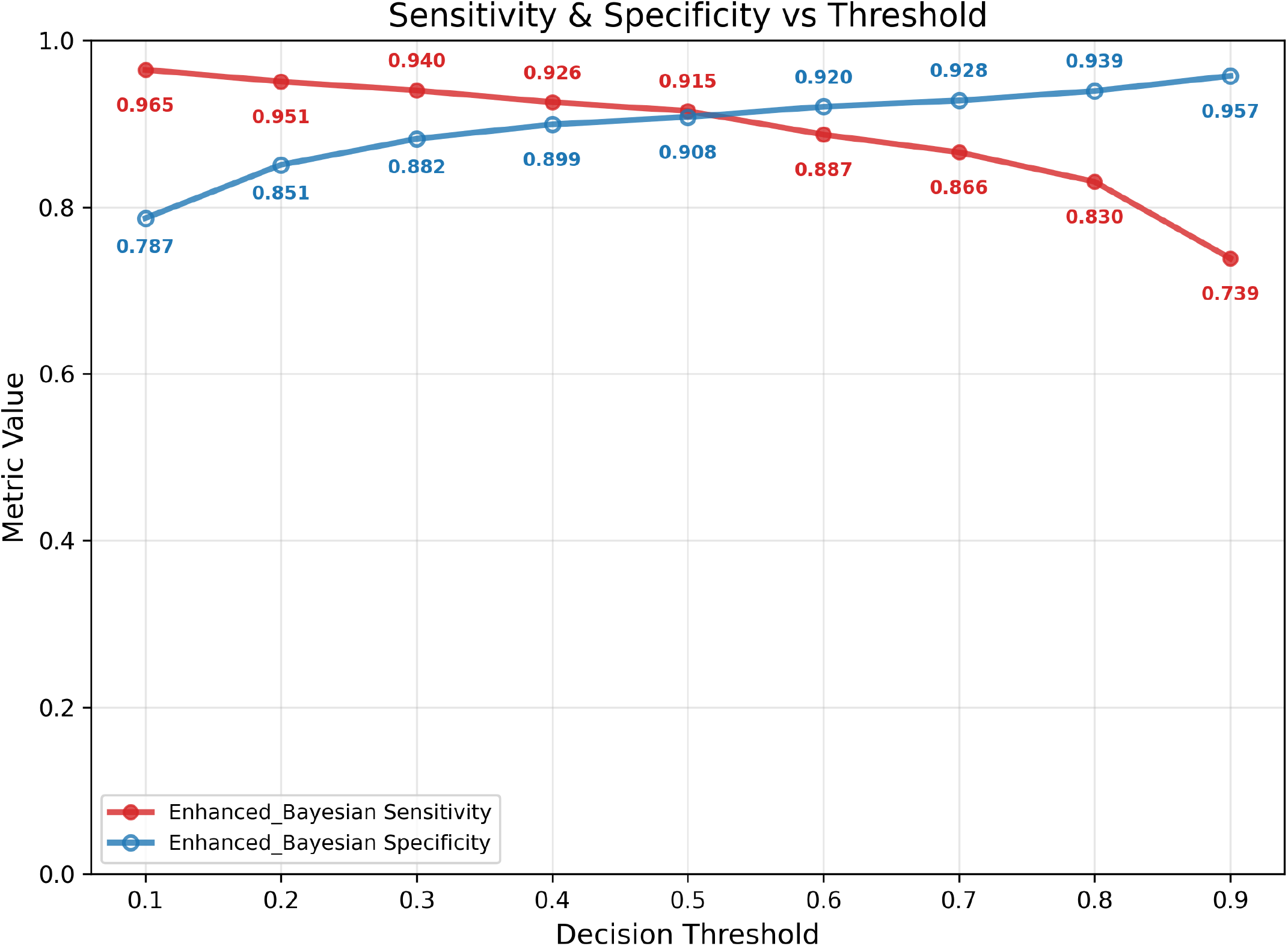
Sensitivity and Specificity across decision thresholds.

### Performance by cancer type

The point-estimate sensitivity was high across all major cancer groups, ranging from 92.0% for head and neck cancers to over 94% for upper and lower gastrointestinal cancers (Table 4). Sensitivity remained *>*87% in the four largest groups (head and neck, breast, upper GI, gynecologic), supporting consistent detection across diverse tumor types. The apparent 100% sensitivities for lung/thoracic, and male genitourinary cancers reflect small sample sizes (n *<* 10) and are accompanied by wide confidence intervals, indicating that these estimates should be interpreted cautiously. The “Others” category, which pooled rare cancers, showed wider variability with a sensitivity of 76.9% (95% CI: 53.8–100.0). The test set sensitivity reached 92.2% for all cancer groups excluding the “Others” category, compared to an overall sensitivity of 91.5% when all groups were included; the controls were not subdivided by cancer group. Taken together, these findings suggest that canine detection performance was robust across the cancer types most prevalent in India (head and neck, gastrointestinal, and breast cancer), without evidence of collapse in rarer categories, although precision in smaller groups will require larger cohorts.

**Table 4:**
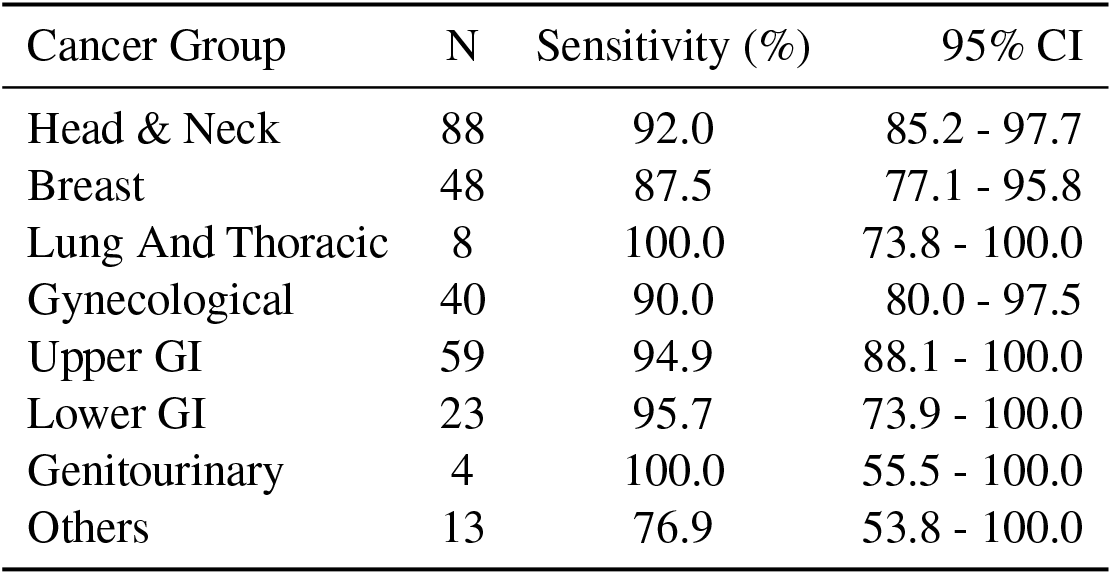
Sensitivity by cancer group in the test cohort.

### Performance by stage

The stage-specific sensitivity remained consistently high across all stages, 87.1% in stage I, 90.8% in stage II, 93.7% in stage III, and 91.5% in stage IV (Table 5). Aggregated across stages, early-stage disease (I–II) was detected with a sensitivity of 89.6%, comparable to 92.5% for late-stage disease (III–IV). Confidence intervals overlapped across stages, and estimates for the “unknown” stage group (n = 14) were imprecise due to the small sample size.

**Table 5:** Sensitivity by cancer stage.

| Stage | N | Sensitivity (%) | 95% CI |
| --- | --- | --- | --- |
| I | 31 | 87.1 | 74.2 - 96.8 |
| II | 65 | 90.8 | 83.1 - 96.9 |
| III | 79 | 93.7 | 88.6 - 98.7 |
| IV | 94 | 91.5 | 85.1 - 96.8 |
| Unknown | 14 | 92.9 | 78.6 - 100.0 |

### Subgroup-analysis

Performance was broadly consistent across the demographic and exposure strata (Table 6). The sensitivity was somewhat higher in participants aged *>*50 years (94.8%) than in those *<*50 years (85.9%), whereas the specificity was higher in younger individuals (95.1% vs 85.0%). Male and Female participants showed comparable sensitivity (90.6% vs 92.2%) and specificity (89.2% vs 91.7%), robust against differences in cancer type distribution (e.g., breast and gynecologic vs head and neck). Importantly, detection performance was resilient in both never-tobacco users (sensitivity 90.6%) and current/former users (92.7%), with overlapping confidence intervals, suggesting that discrimination was not solely attributable to tobacco-related exposures.

**Table 6:**
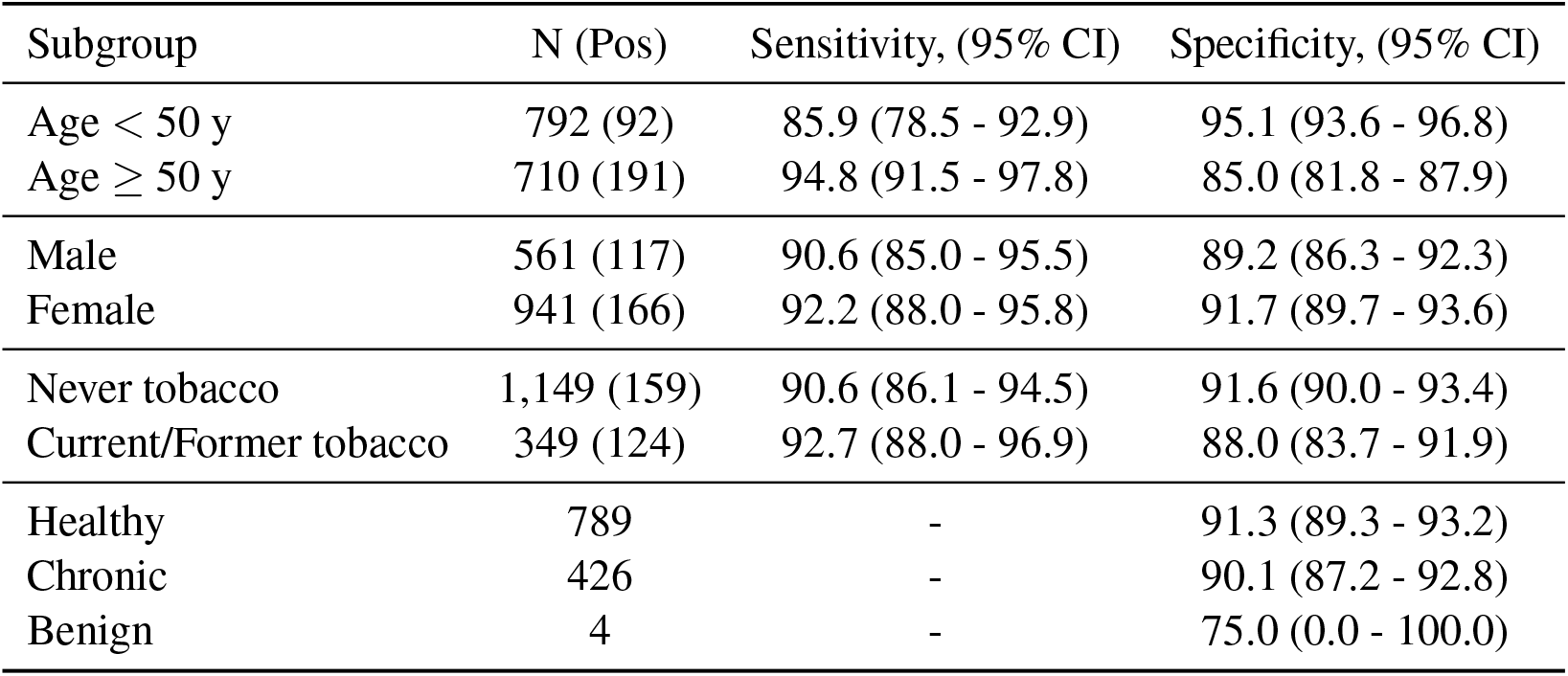
Subgroup performance.

### Robustness to Individual Dog Variation

Although all dogs were trained under an identical protocol, the individual-level sensitivity and specificity varied (Figure S1 and Figure S2). Because such variability can raise concerns regarding replicability and standardization in biomedical detection, we evaluated the robustness of the ensemble using two stress tests.

#### Leave-one-dog-out

We conducted an ablation analysis in which, for each of the seven dogs, the dog was removed from the ensemble and the Bayesian fusion model was re-evaluated without its indications. This procedure yields seven “reduced” ensembles, each reflecting performance if a given dog were unavailable. The results are summarized in Table 7.

**Table 7:** Ablation study: Leave-One-Dog-Out.

| Dog (N) | Individual:<br>Sensitivity,<br>Specificity (%) | (Ensemble)-(dog):<br>Sensitivity,<br>Specificity (%) | (Ensemble)-(dog)<br>AUC-ROC |
| --- | --- | --- | --- |
| Chloe (1460) | 83.6, 83.6 | 91.5, 90.2 | 0.960 |
| Cheetah (1391) | 84.9, 82.2 | 89.0, 90.7 | 0.955 |
| Iggy (699) | 82.1, 84.6 | 91.5, 90.2 | 0.961 |
| Billy (1459) | 82.3, 82.5 | 91.5, 91.1 | 0.957 |
| Whiskey (242) | 88.2, 88.0 | 91.2, 91.2 | 0.962 |
| Jessie (383) | 86.7, 84.8 | 91.2, 91.3 | 0.959 |
| Bane (1251) | 81.0, 85.0 | 87.3, 92.3 | 0.963 |
| Ensemble | 91.5, 90.8 | - | 0.962 |

#### “Bad-day” simulation

To model the transient degradation in one dog’s signal at the session level, we performed a permutation stress test. For each testing session, we randomly selected one active dog and permuted the dog’s indications across the session’s samples, preserving all other dogs’ indications and the ground-truth labels. This destroys the association between the dog’s responses and the case/control status while preserving its marginal response rate. We repeated the procedure five times with distinct random seeds and reported the mean area under the ROC curve (AUC) across the replicates (Fig. 4).

**Figure 4.**
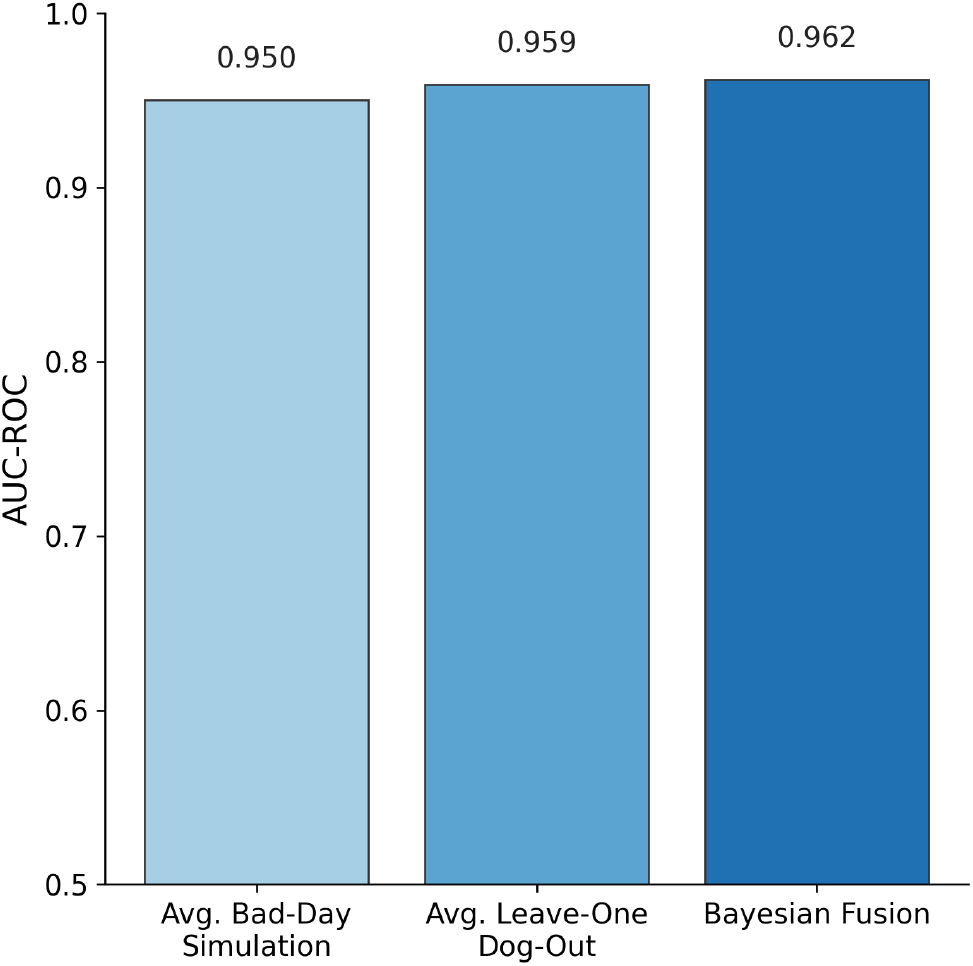
Robustness of our model against individual dog variation.

#### Interpretation

Across both analyses, ensemble performance remained stable relative to the full model, indicating that the fusion approach is resilient to (i) the removal of any single dog and (ii) temporary within-session degradation in one dog’s indications. This supports the robustness of multi-dog Bayesian fusion to idiosyncratic behavior at the individual-dog level.

### EHR-Derived Prior Contribution

We quantified the relative contribution of electronic health record (EHR)-derived covariates to the prior *P*_0_ using standardized contribution scores (percent importance, averaged across crossvalidated folds). The largest effects were observed for tobacco exposure (chewing+smoking) (29.50%), persistent symptoms (14.45%) and age group (8.02%), followed by sex (6.33%), first-degree family history of cancer (6.28%) and comorbidities (5.32%)(Table 8). The directions and relative magnitudes align with established epidemiology — such as risk increasing with age and cumulative tobacco exposure and first-degree family history elevatating baseline risk. Incorporating these predictors (including the composite tobacco measure and an age*×*medication interaction) improved discrimination of the prior model by ΔAUC = +0.11 versus reduced specifications. Contribution ranks were stable under resampling, supporting the face validity and robustness of the priors and their added value when fused with canine indications.

**Table 8:** Prior contribution.

| Table 8: Prior contribution |  |
| --- | --- |
| Variable | Contribution (%) |
| Tobacco - chewing | 23.74 |
| Persistent Symptoms | 14.45 |
| Age Group | 8.02 |
| Sex | 6.33 |
| Family Cancer History | 6.28 |
| Tobacco - smoking | 5.76 |
| Comorbidities | 5.32 |

### Ablation: Contribution of model components

Figure 5 compares three operating variants on the test set: (i) *Only Prior*, which uses the EHR–derived covariate model without dog indications; (ii) *Pack Majority*, an unweighted majority vote across dogs that ignores covariates; and (iii) full *Bayesian Fusion*, which combines per-dog likelihoods with the prior. Discrimination improved from an AUC of 0.832 (Only Prior) to 0.932 with Pack Majority (Δ= + 0.100), and further to 0.962 with Bayesian Fusion (Δ= + 0.030 vs. majority vote; Δ= + 0.130 vs. prior). These results indicate that canine indications provide the dominant signal, whereas covariate-informed prior adds incremental, complementary information when fused probabilistically, thereby yielding a modest but meaningful gain over naive voting and producing well-defined posterior probabilities for thresholding.

**Figure 5.**
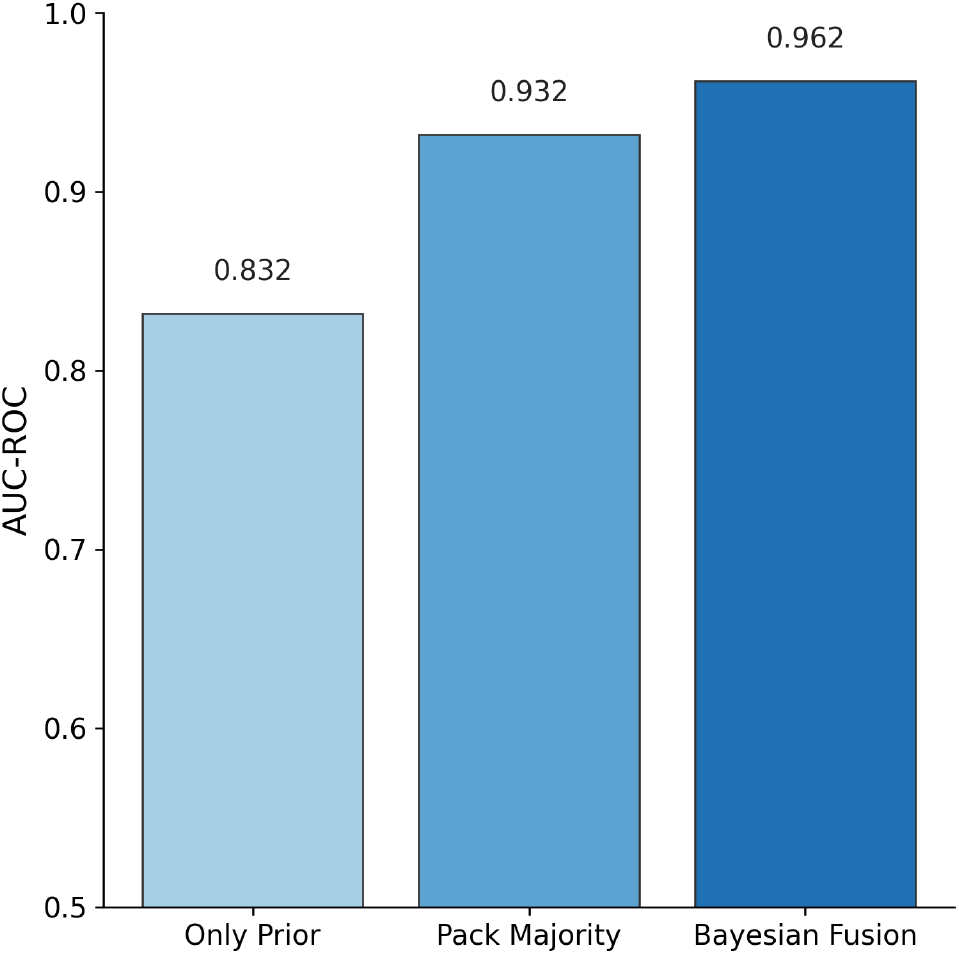
Ablation : Performance of Model Components.

## Discussion

India’s National Program for Prevention and Control of Non-Communicable Diseases (NP-NCD) operationalizes population-based screening for three cancers, oral, cervical, and breast, delivered through Health and Wellness Centers by healthcare workers using oral visual examination, VIA, and clinical breast examination, respectively.^23^ Despite national scale-up, screening coverage remains modest,^24^ and these three cancers together account for only about one-third of the incident cancers in India.^3^ In low-prevalence populations, even highly specific tests yield a modest positive predictive value (PPV), amplifying the downstream workload.^8^ A non-invasive, acceptable, low-cost triage test at the front end of NP-NCD pathways could enrich pretest probability and focus confirmatory diagnostics on a smaller, higher-risk subset—an approach aligned with the CanTest framework for evaluating tests across real-world diagnostic pathways.^9^

Against this backdrop, we evaluated the analytical validity of multi-cancer breath-based canine detection. In a multi-center, assessor-masked case–control study across six Indian hospitals, a Canine–Bayesian fusion model demonstrated strong analytical validity (ROC AUC = 0.962) with balanced sensitivity and specificity above 90%. Sensitivity remained above 87% across stages I to IV, and performance was broadly consistent across major cancer groups, with the Bayesian fusion model mitigating between-dog variability relative to individual reads. These data support the potential role of breath-based triage in expanding participation and improving the efficiency of downstream diagnostics within workflows such as NP-NCD guidelines. However, prospective, population-based studies are needed to calibrate operating thresholds and quantify clinical utility under true screening prevalence.

Breath-based detection interrogates volatile metabolic signatures and is therefore mechanistically complementary to blood-based multi-cancer early detection (MCED) assays that rely on circulating genomic/epigenomic markers. Our results suggest integrated pathways that can be evaluated across settings: *(i)* breath-first triage to enrich pretest probability before organ-specific imaging or blood-based MCED; *(ii)* reflex testing (breath → MCED or MCED → breath) to improve PPV while controlling downstream workload; and *(iii)* probabilistic fusion of breath and blood signals for a single operating point tailored to local resources. Such designs could increase reach (primary care, workplace, or home-based breath collection), improve acceptability, and reduce false-positive results. Head-to-head and combined-pathway studies—incorporating cost, throughput, and qualitycontrol metrics will be critical for determining where breath-based triage adds the greatest value in both high- and low-resource health systems.

However, important limitations of this study remain. First, the case–control design is susceptible to spectrum and selection biases and typically inflates apparent performance relative to true screening settings. Second, the cancer prevalence in the test cohort (*∼*1:4) exceeds that of the general screening population, which would depress the PPV at low prevalence despite high sensitivity. Third, generalizability beyond participating centers cannot be stated with certainty, all sites were within a single Indian state (Karnataka), and unmeasured geographic, environmental, dietary, or microbiome factors may shift breath VOC backgrounds. Finally, batch effects and residual confounding were possible despite standardized procedures.

### Planned methodological enhancements

To further reduce between-dog variability and strengthen the data-driven framework, we plan to augment the fusion with (i) computer-vision features from multi-view videos to estimate features such as dwell time, head pose, re-approach counts, and micro-pauses extracted via pose-estimation and event-detection models, and (ii) non-invasive physiological signals, including wearable EEG to capture brain activity before and during sniffing. In a hierarchical Bayesian formulation, dog-specific likelihood terms can be dynamically weighted by these signals and by contemporaneous performance calibration, yielding time-varying reliability weights for each dog–trial. This refinement is expected to narrow the uncertainty around the persample posterior probabilities, attenuate variable behavior, improve repeatability across sessions and sites, and improve the overall performance of the system.

### Prospective evaluation

These results establish analytical validity but require confirmation in prospective, population-based studies to determine the clinical utility of true screening prevalence. Such studies should pre-specify thresholds and referral pathways and evaluate key endpoints (positive predictive value, detection rate per 1,000 screened, stage distribution at diagnosis, and time to diagnostic resolution) while monitoring downstream diagnostic yield, false-positive cascades, and participant acceptability. Integration within standard-of-care pathways and assessments of throughput, workflow, and costs will be essential but are beyond the scope of the present case–control study.

In summary, breath-based multi-cancer detection using trained dogs augmented by Bayesian fusion demonstrated a strong analytical performance across cancer types and stages. The next phase of methodologically enriched modeling with EEG and machine vision, along with prospective evaluation and health-system integration, will be critical for determining clinical utility and implementation feasibility.

## Data Availability

All data produced in the present study are available upon reasonable request to the authors

## Acknowledgments

We are deeply grateful to all patients who volunteered and provided breath samples for this study.

We gratefully acknowledge the contributions of clinical partners across all sites involved in the BreathEasy study. At Radon Cancer Center, Hubli, we thank Dr. Sheetal Kulgod for leading the screening camps and Dr. Vishal Malvade for patient recruitment support. At KMCRI, Hubli, we thank Dr. Laxmikant, Dr. Manisha,, Dr Vishal Kulkarni, Dr Sunita Vernekar, Dr Rajesh H, Dr Parvati Jugular, Dr Kanchana, Nurse Ashwini, Manjunath and Chandrashekar for their support in recruitment. At KCTRI, Hubli, we acknowledge Dr. Umesh Hallikeri and Dr. Vilas Gunda for facilitating site access and screening camps, along with Dr. Saikumari and Dr. Poornachandra for their contributions to participant recruitment. We also extend our thanks to Dr. Nikhil, Bhagya and Janaki Sisters at St. John’s Medical College, Bangalore; Dr. Uday Shivaji, Dr. Karthik, Dr Anjum and Veena at Narayana Health City, Bangalore; and Mr. Surinder Kher and Ms. Rajeshwari at Aster CMI Hospital, Bangalore for site-level clinical operations support. We also thank Dr Shashikant Kulgod and Dr Akash Gadgade for their support throughout the study.

We sincerely thank our Clinical Research Coordinators for their on-ground efforts and dedication to sample and data collection : Kiran B Nagashettikoppa, Rudresh Hiremath, Dr. Arnab Sarkar, Dr. Basavaraj Varad, Akash Sudi, Dr. Tahameen Kurubanavar, Dr. Anjum Jangliwale, Dasmen Pratiban, Dr. Kishore and Dr. Shabin. We also thank Dr Minal Dakhave for her support throughout her tenure.

We thank the Dognosis lab team — Richard King, Manish, Gokul, Nirmal, and Venkat Rao — for their dedicated work managing canine operations.

We recognize the contributions of the Dognosis R&D team, including Aswin R, Thirumalai Srinivasan, Abhijit Kadalli, Vishnu Kumar, Sree Subha Ramaswamy, Dilawar Singh, Maadesh Kumar, Subham Hota, Achin Parashar, Anushree Narjala, Shashikant Gadiwaddar, and Promit Moitra for their efforts in hardware development, data management, analysis, and model building.

This study was supported by Dognosis India Pvt. Ltd.

## Author Contributions

Conception and design: Akash Kulgod, Itamar Bitan, Sanjeev Kulgod, Sahana Shanbhag, Claire Guest, Rob Harris

Provision of study materials or patients: Sanjeev Kulgod, Basavaraj R. Patil, Shashidhar K., Rakesh S. Ramesh, Kiran Kulkarni, Somashekhar S. P., Swaratika Majumdar, Akshita Singh

Collection and assembly of data: Sahana Shanbhag, Ido Aviram, Itamar Bitan, Akash Kulgod

Data analysis and interpretation: Akash Kulgod, Itamar Bitan, Sahana Shanbhag, Ido Aviram

Manuscript writing: Akash Kulgod (drafting); all authors (review and editing)

Final approval of manuscript: All authors

All authors are accountable for all aspects of the work

## Authors’ Disclosures of Potential Conflicts of Interest

The following represents the disclosure of information provided by the authors of this manuscript. All relationships were considered compensated unless otherwise noted. Relationships are self-held unless otherwise noted. I = Immediate Family Member, Inst = My Institution.

Relationships may not be related to the subject matter of this study.

- **Sanjeev Kulgod, MCh** Employment: RadOn Cancer Centre, Hubli. Role: Lead Principal Investigator. No other conflicts to disclose
- **Basavaraj R. Patil, MS, FRCS** Employment: Karnataka Cancer Therapy and Research Institute, Hubli. No other conflicts to disclose .
- **Shashidhar K**., **MCh** Employment: Karnataka Medical Centre and Research Institute, Hubli. No other conflicts to disclose.
- **Rakesh S. Ramesh, DNB** Employment: St John’s Medical College, Bangalore. Research Funding: Dognosis India Pvt. Ltd (Inst).
- **Kiran Kulkarni, MCh** Employment: St John’s Medical College, Bangalore. Research Funding: Dognosis India Pvt. Ltd (Inst).
- **Somashekhar S. P**., **MCh** Employment: Aster CMI Hospital, Bangalore. Research Funding: Dognosis India Pvt. Ltd (Inst).
- **Swaratika Majumdar, DM** Employment: Narayana Health City, Bangalore. No other conflicts to disclose .
- **Akshita Singh, DNB** Employment: Narayana Health City, Bangalore. No other conflicts to disclose .
- **Claire Guest** Employment: Medical Detection Dogs. No other conflicts to disclose .
- **Rob Harris** Employment: Medical Detection Dogs. No other conflicts to disclose .
- **Sahana Shanbhag** Employment: Dognosis India Pvt. Ltd. Stock and Other Ownership Interests: Dognosis Inc (ESOPs).
- **Ido Aviram** Employment: Dognosis India Pvt. Ltd. Stock and Other Ownership Interests: Dognosis Inc (ESOPs).
- **Itamar Bitan** Employment: Dognosis India Pvt. Ltd. Stock and Other Ownership Interests: Dognosis Inc.
- **Akash Kulgod** Employment: Dognosis India Pvt. Ltd. Stock and Other Ownership Interests: Dognosis Inc.

## Supplementary Material

**Table S1:**
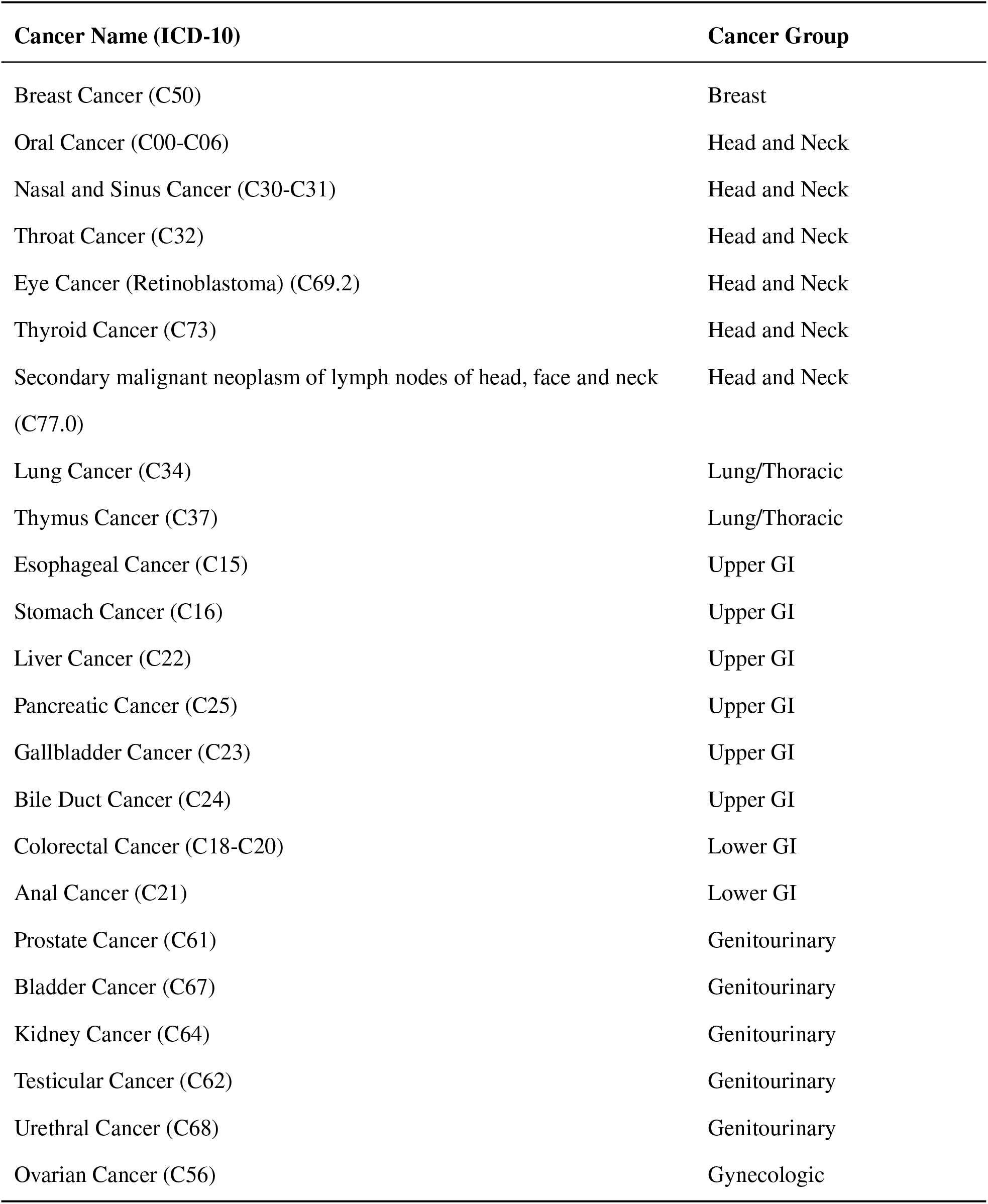
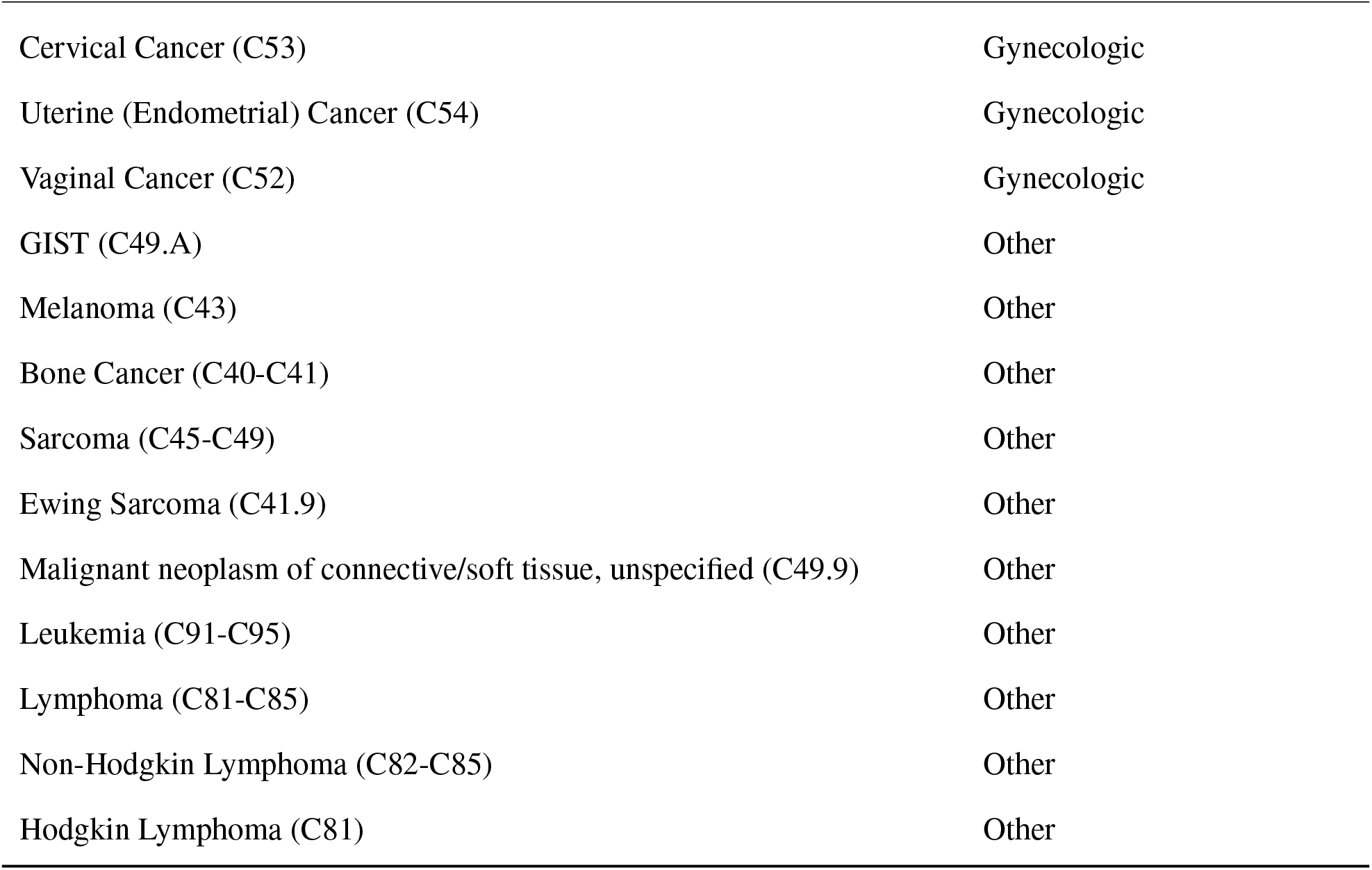
Categorization of cancer types.

**Table S2:** Baseline characteristics, training cohort (n=1,773)

| Characteristic | Cancer (n=373) | Non-cancer (n=1,400) | Total (n=1,773) |
| --- | --- | --- | --- |
| Male, n (%) | 172 (46.1) | 699 (49.9) | 871 (49.1) |
| Female, n (%) | 201 (54.0) | 701 (50.1) | 902 (50.9) |
| Age, mean (SD) | 55.2 (13.2) | 46.6 (14.1) | 48.4 (14.3) |
| **Tobacco (smoking)** |  |  |  |
| Ever, n (%) | 41 (11.0) | 121 (8.6) | 162 (9.1) |
| Current, n (%) | 5 (1.3) | 53 (3.8) | 58 (3.3) |
| Past, n (%) | 36 (9.7) | 68 (4.9) | 104 (5.9) |
| Never, n (%) | 331 (88.7) | 1,277 (91.2) | 1,608 (90.7) |
| **Tobacco (chewing)** |  |  |  |
| Ever, n (%) | 119 (31.9) | 244 (17.4) | 363 (20.4) |
| Current, n (%) | 10 (2.7) | 167 (11.9) | 177 (10.0) |
| Past, n (%) | 109 (29.2) | 77 (5.5) | 186 (10.5) |
| Never, n (%) | 251 (67.5) | 1,155 (82.5) | 1,406 (79.3) |
| **Comorbidities** |  |  |  |
| Hypertension, n (%) | 82 (22.0) | 280 (20.0) | 362 (20.4) |
| Diabetes, n (%) | 57 (15.3) | 212 (15.2) | 269 (15.2) |
| Other Heart Diseases (Other CVD), n (%) | 2 (0.5) | 59 (4.2) | 61 (3.4) |
| Kidney Disease, n (%) | 0 (0.0) | 33 (2.4) | 33 (1.9) |
| Ischaemic Heart Disease (IHD), n (%) | 5 (1.3) | 12 (0.9) | 17 (1.0) |

**Table S3:** Cancer-group distribution in the training cohort (n=373)

| Cancer group | n | % of cancers |
| --- | --- | --- |
| Head and Neck | 117 | 31.4 |
| Lung/Thoracic | 12 | 3.2 |
| Breast | 69 | 18.5 |
| Upper GI | 59 | 15.8 |
| Lower GI | 11 | 2.9 |
| Gynecologic | 62 | 16.6 |
| Genitourinary (male) | 18 | 4.8 |
| Other | 25 | 6.7 |
| Total | 373 | 100.0 |

**Table S4:** Distribution of samples across locations in training and test cohorts.

| Location | Training |  | Test |  | Total |  |
| --- | --- | --- | --- | --- | --- | --- |
|  | Cancer | Non-cancer | Cancer | Non-cancer | Training | Test |
| Aster CMI, n (%) | 4 (1.1) | 21 (1.5) | 5 (1.8) | 7 (0.6) | 25 (1.4) | 12 (0.8) |
| Community Camps, n (%) | 2 (0.5) | 205 (14.6) | 0 (0.0) | 727 (59.6) | 207 (11.7) | 727 (48.4) |
| KCTRI, n (%) | 108 (29.0) | 353 (25.2) | 118 (41.7) | 25 (2.1) | 461 (26.0) | 143 (9.5) |
| KMCRI, n (%) | 121 (32.4) | 416 (29.7) | 91 (32.2) | 368 (30.2) | 537 (30.3) | 459 (30.6) |
| Narayana Health City, n (%) | 26 (7.0) | 99 (7.1) | 1 (0.4) | 10 (0.8) | 125 (7.1) | 11 (0.7) |
| RadOn, n (%) | 64 (17.2) | 167 (11.9) | 61 (21.6) | 71 (5.8) | 231 (13.0) | 132 (8.8) |
| ST. JOHNS, n (%) | 48 (12.9) | 139 (9.9) | 7 (2.5) | 11 (0.9) | 187 (10.5) | 18 (1.2) |
| Total | 373 | 1400 | 283 | 1219 | 1773 | 1502 |

**Table S5:** Distribution of comorbid conditions in training and test cohorts.

| Condition | Training |  | Test |  |
| --- | --- | --- | --- | --- |
|  | Cancer | Non-cancer | Cancer | Non-cancer |
| No Condition, n (%) | 256 (68.6) | 939 (67.1) | 180 (63.6) | 790 (64.8) |
| Hypertension, n (%) | 82 (22.0) | 280 (20.0) | 72 (25.4) | 289 (23.7) |
| Diabetes, n (%) | 57 (15.3) | 212 (15.1) | 47 (16.6) | 193 (15.8) |
| Other Heart Diseases (Other CVD), n (%) | 2 (0.5) | 59 (4.2) | 1 (0.4) | 51 (4.2) |
| Kidney Disease, n (%) | 0 (0.0) | 33 (2.4) | 0 (0.0) | 12 (1.0) |
| Ischaemic Heart Disease (IHD), n (%) | 5 (1.3) | 12 (0.9) | 2 (0.7) | 3 (0.2) |
| Epilepsy, n (%) | 0 (0.0) | 3 (0.2) | 0 (0.0) | 15 (1.2) |
| Anemia, Low Iron, n (%) | 1 (0.3) | 12 (0.9) | 1 (0.4) | 2 (0.2) |
| Chronic Obstructive Pulmonary Disease, n (%) | 3 (0.8) | 11 (0.8) | 0 (0.0) | 1 (0.1) |
| Asthma, n (%) | 1 (0.3) | 1 (0.1) | 3 (1.1) | 9 (0.7) |
| Thyroid Disease, n (%) | 1 (0.3) | 7 (0.5) | 1 (0.4) | 0 (0.0) |
| Hernia, n (%) | 1 (0.3) | 4 (0.3) | 0 (0.0) | 3 (0.2) |
| Hemorrhoids, n (%) | 0 (0.0) | 2 (0.1) | 1 (0.4) | 5 (0.4) |
| Liver Disease (Other), n (%) | 0 (0.0) | 4 (0.3) | 2 (0.7) | 2 (0.2) |
| Hepatitis, n (%) | 1 (0.3) | 3 (0.2) | 1 (0.4) | 2 (0.2) |
| Human immunodeficiency virus [HIV] disease, n (%) | 2 (0.5) | 1 (0.1) | 3 (1.1) | 1 (0.1) |
| Pancreatitis, n (%) | 0 (0.0) | 2 (0.1) | 0 (0.0) | 4 (0.3) |
| Mental and behavioural (psychiatric) disorders, n (%) | 0 (0.0) | 1 (0.1) | 0 (0.0) | 5 (0.4) |
| Urinary Tract Infection, n (%) | 0 (0.0) | 3 (0.2) | 2 (0.7) | 0 (0.0) |
| Acute appendicitis, n (%) | 0 (0.0) | 4 (0.3) | 0 (0.0) | 1 (0.1) |

**Table S6:** Specificity & Sensitivity by location of the sample.

| Location | N (Positive) | Sensitivity (%) | Specificity (%) |
| --- | --- | --- | --- |
| Aster CMI | 12 (5) | 80.0 | 100.0 |
| Community Camps | 727 (0) | - | 96.3 |
| KCTRI | 143 (118) | 97.5 | 56.0 |
| KMCRI | 459 (91) | 83.5 | 85.9 |
| Narayana Health City | 11 (1) | 100.0 | 100.0 |
| RadOn | 132 (61) | 91.8 | 73.2 |
| ST. JOHNS | 18 (7) | 100.0 | 72.7 |

**Figure S1:**
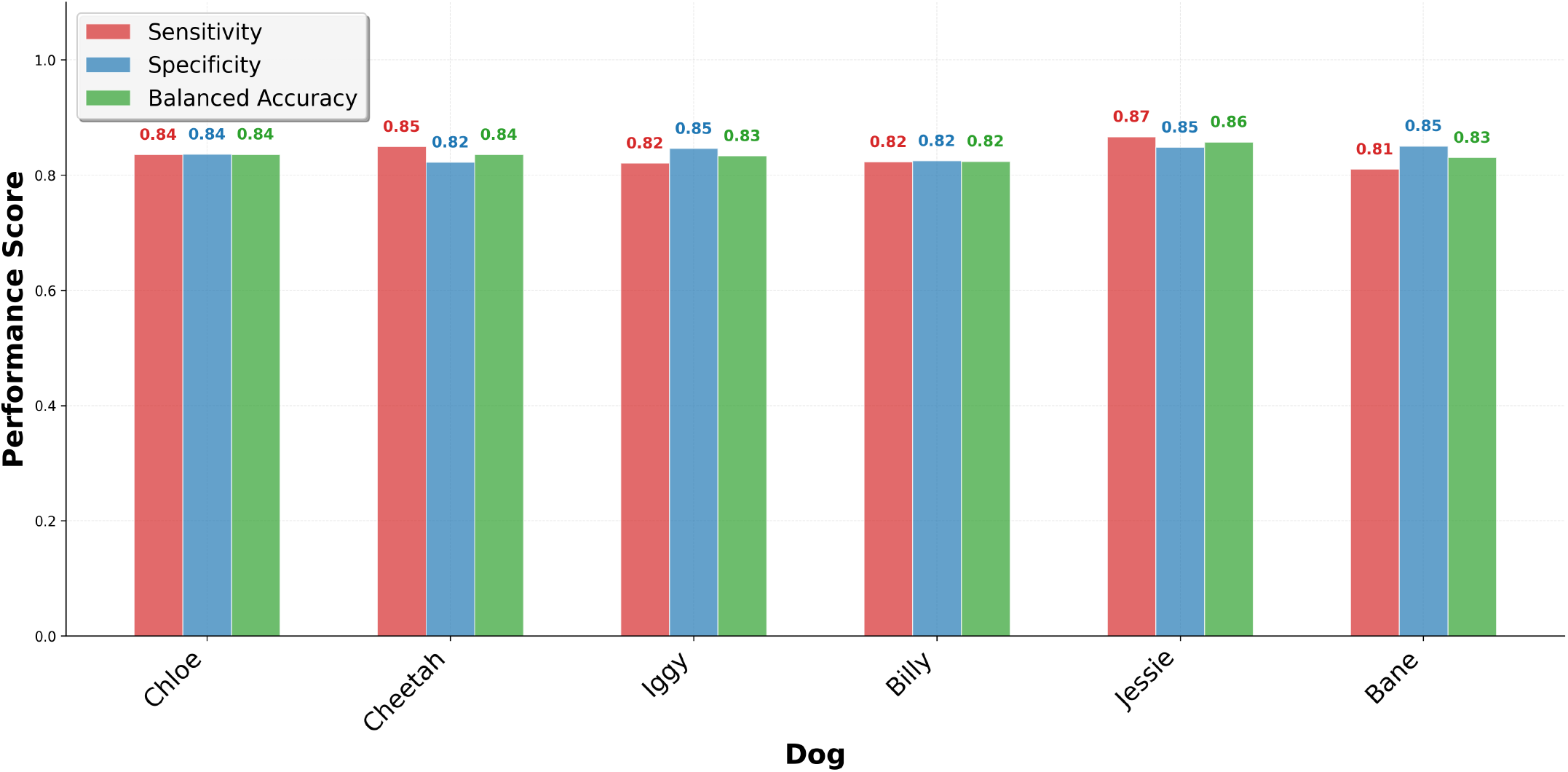
Sensitivity and specificity for each dog.

**Figure S2:**
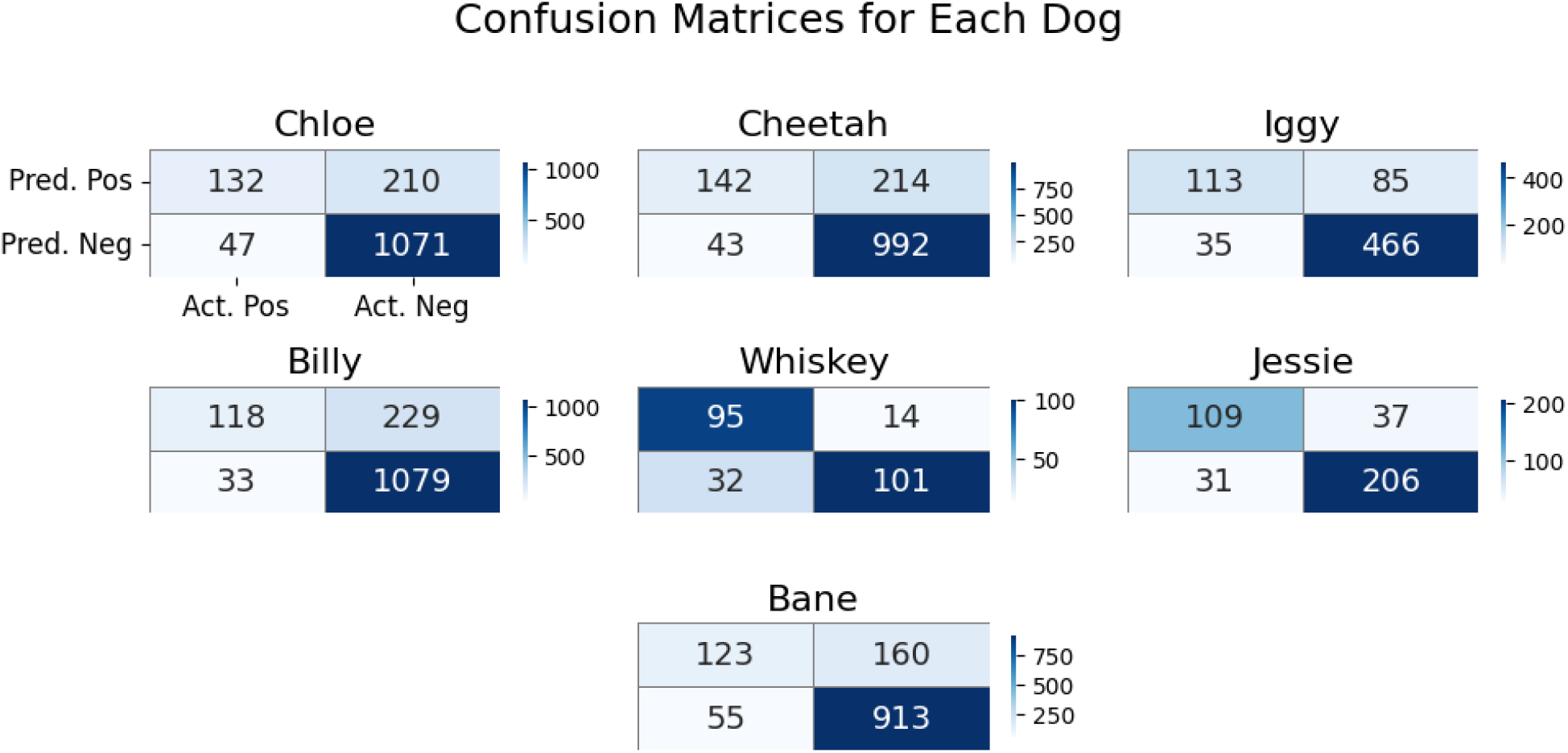
Confusion matrices for each dog.

## Notes

### Funding Statement

This study was funded by Dognosis India Pvt Ltd

### Author Declarations

Ethics Committee of Karnataka Medical College and Research Institute, Hubli, gave ethical approval for this work. Ethics Committee of St Johns Medical College, Bangalore, gave ethical approval for this work. Institutional Ethics Committee of Aster CMI Hospital, Bangalore, gave ethical approval for this work. Institutional Ethics Committee of Narayana Health City, Bangalore, gave ethical approval for this work.

